# A Fast and Interpretable Logistic Regression Framework for Breast Tumor Classification Using the Wisconsin Diagnostic Dataset

**DOI:** 10.64898/2025.12.23.25342946

**Authors:** Weihao Cheng, Zekai Yu

## Abstract

Early and reliable discrimination between malignant and benign breast tumors is essential for clinical decision-making and for reducing unnecessary invasive procedures. This study presents a lightweight and reproducible machine-learning pipeline that integrates standard feature normalization with logistic regression to classify breast tumors using the Breast Cancer Wisconsin (Diagnostic) dataset (WDBC), which contains 569 samples described by 30 quantitative features derived from digitized fine-needle aspirate (FNA) images [1–3]. We implemented an end-to-end workflow in Python (scikit-learn), including stratified train–test splitting, model training, and evaluation with clinically meaningful metrics such as accuracy, sensitivity, specificity, and ROC-AUC [4–6]. On the held-out test set (n=114), the proposed approach achieved 98.25% accuracy and a ROC-AUC of 0.9954, with a confusion matrix indicating only two misclassifications (1 malignant predicted as benign, 1 benign predicted as malignant). Specifically, when treating malignant cases as the clinically critical positive class, the method yielded 97.62% sensitivity and 98.61% specificity. These results demonstrate that a simple, interpretable model can achieve near state-of-the-art performance on structured biomedical features while remaining computationally efficient and suitable for rapid prototyping.In addition, we benchmarked logistic regression against several classical baselines (SVM with RBF kernel, random forest, and kNN) under the same train–test split and evaluation protocol. The results indicate that increasing model complexity yields limited performance gains on WDBC, suggesting that an interpretable linear classifier can already approach the performance ceiling on this feature-engineered dataset. Future work will focus on external validation, calibration for risk estimation, and multimodal extensions to incorporate imaging or omics signals.

## 1. Introduction

Breast cancer remains a major global health burden, and timely diagnosis is closely tied to improved outcomes. In clinical workflows, suspicious breast lesions are often evaluated using imaging and cytology/histopathology. However, borderline presentations and inter-observer variability can lead to uncertainty and potentially unnecessary biopsies or delayed intervention. Consequently, computer-aided diagnosis (CAD) and machine-learning (ML) techniques have been widely explored to support clinicians by offering consistent and quantitative decision support.

From a clinical perspective, decision support tools are most valuable when they can help clinicians manage uncertainty at the margin—particularly for lesions that do not present with clear-cut malignant characteristics. In such scenarios, a model that provides calibrated risk scores and interpretable evidence (e.g., which morphological properties drive the prediction) can assist clinicians in deciding whether to recommend further diagnostic procedures, short-interval follow-up, or biopsy. Therefore, interpretability and reliability are not merely “nice-to-have” properties, but practical requirements for clinical adoption.

From an engineering perspective, ML-based decision systems for healthcare must satisfy several practical requirements: (i) strong discriminative performance, (ii) transparency/interpretability to build clinical trust, (iii) minimal computational cost for deployment, and (iv) reproducibility for validation and regulatory scrutiny. While deep learning has achieved impressive performance in many imaging tasks, simpler classical ML models remain highly competitive when the input consists of well-engineered features and the dataset size is moderate, as is typical for many clinical structured datasets.

In many real-world medical engineering settings, researchers and clinicians often face constraints that differ from those assumed in large-scale deep learning studies: limited sample sizes, heterogeneous data acquisition processes, and restricted compute budgets. Under these conditions, classical machine learning models—when paired with informative, domain-driven features—can provide strong and reproducible baselines. Such baselines are crucial not only for rapid prototyping, but also for establishing trustworthy reference points before investing in more complex modeling strategies.

In this work, we target a fast-to-implement yet clinically meaningful benchmark: binary classification of breast tumors using the Wisconsin Diagnostic Breast Cancer dataset (WDBC) hosted by the UCI Machine Learning Repository [1]. The WDBC dataset includes 569 cases with 30 real-valued features computed from digitized images of FNA samples, and it has served as a foundational benchmark for ML-based tumor classification [2,3]. We propose a streamlined pipeline using standardization + logistic regression, emphasizing engineering simplicity, interpretability, and strong baseline performance. We also provide a clear experimental protocol and evaluation outputs (ROC curve and confusion matrix) that are easy to include in a short academic paper.

In addition, we conduct comparative experiments with several classical machine-learning baselines to contextualize the performance of logistic regression within a broader modeling landscape.

## 2. Related Work

The application of machine learning to breast cancer diagnosis has a long history, particularly in the context of cytological and morphological feature analysis. One of the most influential early efforts was introduced by Street, Wolberg, and Mangasarian, who proposed quantitative nuclear feature extraction from digitized fine-needle aspirate (FNA) images and demonstrated that these features could effectively discriminate between malignant and benign breast tumors [2]. Their work laid the foundation for feature-based machine learning approaches in breast cancer diagnosis and highlighted the clinical relevance of nuclear morphology.

Building upon this foundation, Mangasarian et al. further explored optimization-based learning methods for breast cancer diagnosis and prognosis, employing linear programming techniques to achieve strong classification performance while maintaining interpretability [3]. These studies collectively established that carefully engineered morphological features can encode meaningful diagnostic information, enabling accurate classification even with relatively simple models.

Logistic regression (LR) has remained a widely adopted method in medical and biomedical research due to its probabilistic formulation, interpretability, and robustness on small-to-medium sized datasets. In clinical statistics and epidemiology, LR is often preferred because model coefficients can be directly interpreted as indicators of risk contribution, facilitating communication between data scientists and clinicians [7]. When combined with appropriate feature normalization, LR has been shown to achieve competitive performance across a range of structured clinical prediction tasks.

In contrast to traditional machine learning approaches, recent years have seen rapid growth in deep learning-based methods for breast cancer analysis, particularly convolutional neural networks (CNNs) applied to mammography, ultrasound, and histopathological whole-slide images. These models have demonstrated impressive performance in large-scale imaging datasets but typically require substantial amounts of labeled data, computational resources, and careful regularization to avoid overfitting. Moreover, their complex architectures often limit transparency, making clinical interpretation and regulatory approval more challenging.

As a result, there has been renewed interest in interpretable and trustworthy artificial intelligence for medical applications. Explainable machine learning models, including linear models and tree-based methods, are increasingly recognized as valuable tools when data availability is limited or when model transparency is a primary concern. In such settings, simpler models can provide strong baselines while offering clearer insight into decision mechanisms, particularly when feature design incorporates domain knowledge.

For model evaluation, receiver operating characteristic (ROC) analysis and the area under the ROC curve (AUC) are widely accepted standards for assessing diagnostic performance. These metrics allow threshold-independent comparison of classifiers and are especially useful in medical contexts where sensitivity and specificity trade-offs must be adjusted according to clinical priorities. The nonparametric method proposed by DeLong et al. remains a cornerstone for statistically grounded ROC analysis in biomedical research [5].

In this context, the present study positions itself as a lightweight, interpretable baseline that bridges classical statistical learning and modern medical AI practice. By focusing on reproducibility, interpretability, and engineering simplicity, our work complements existing deep learning approaches and provides a transparent reference point for future methodological development and clinical translation.

## 3. Methods

### 3.1 Dataset

We used the Breast Cancer Wisconsin (Diagnostic) dataset (WDBC) from the UCI Machine Learning Repository [1]. The dataset contains 569 samples and 30 continuous features computed from digitized FNA images. Each sample is labeled as malignant or benign. The UCI repository provides standardized metadata and a persistent DOI, facilitating reproducibility [1]. Prior studies have documented the dataset’s clinical context and feature construction, linking quantitative nuclear morphology to diagnostic outcomes [2,3].

The class distribution of the dataset is illustrated in Figure 1, showing a moderate imbalance with a higher proportion of benign cases, which motivates the use of stratified sampling during train–test splitting.

**Figure 1.**
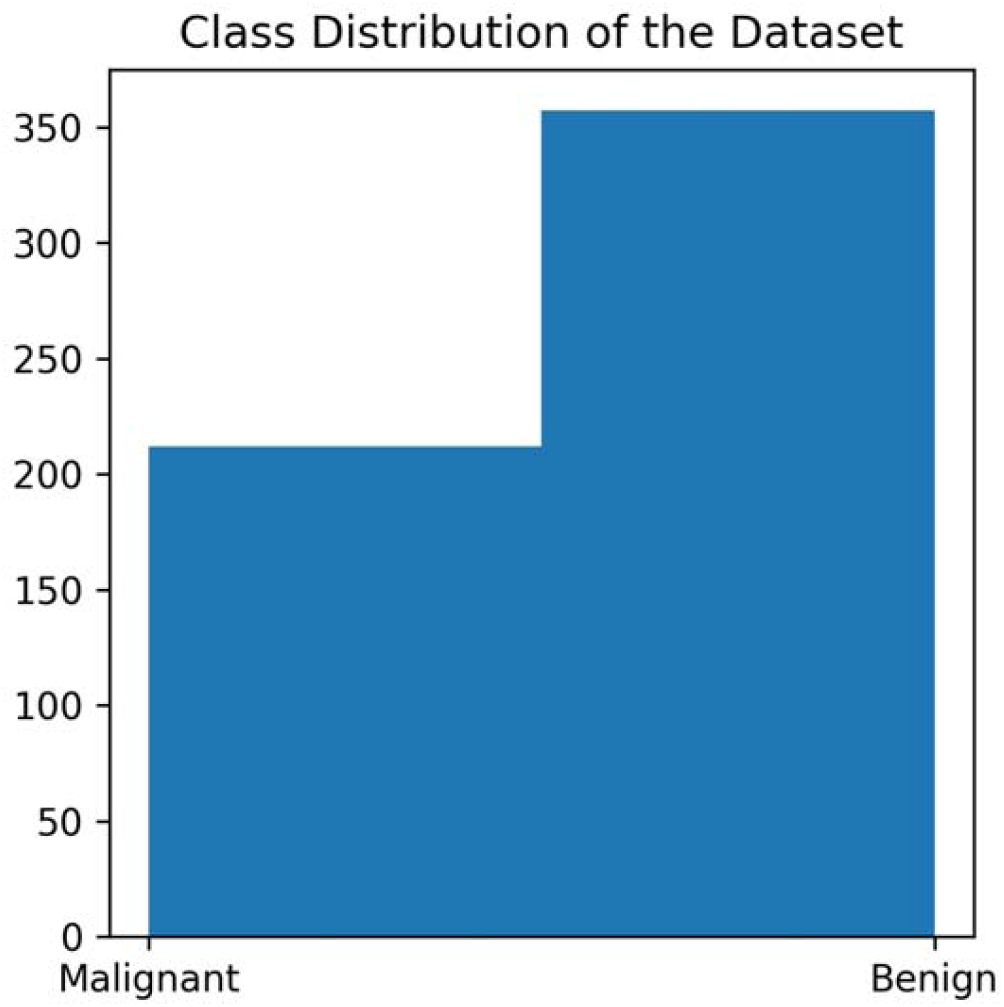
Class distribution of the dataset

**Figure 2.**
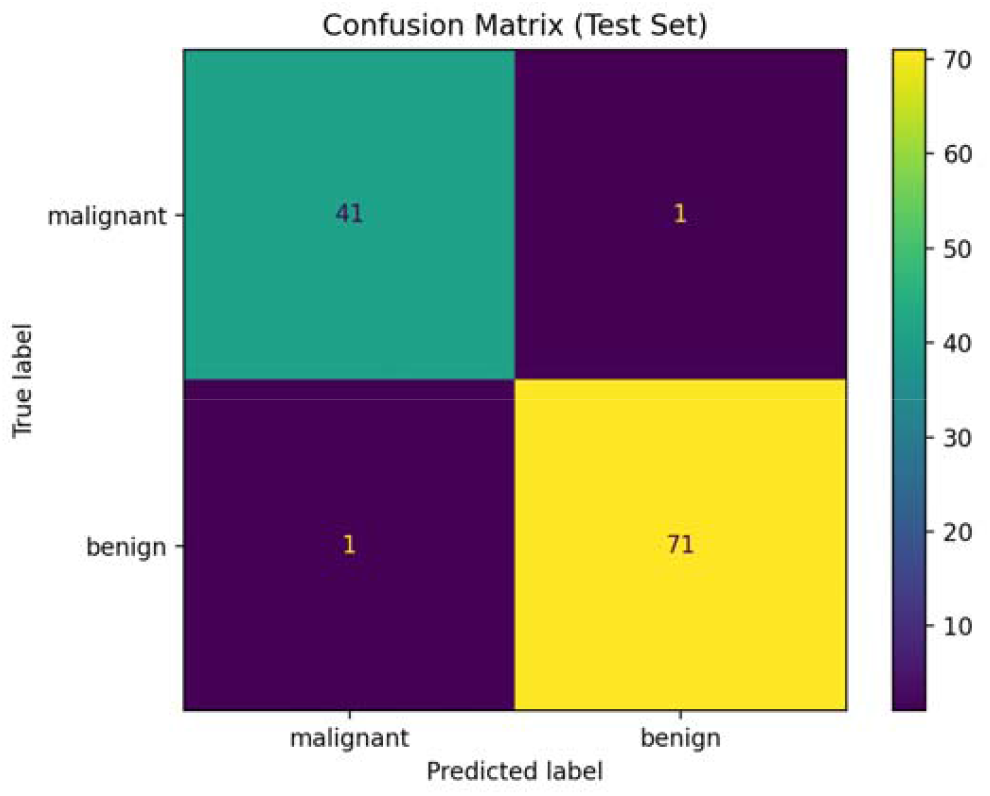
Confusion matrix on the held-out test set, showing near-perfect discrimination with only two misclassifications.

**Figure 3.**
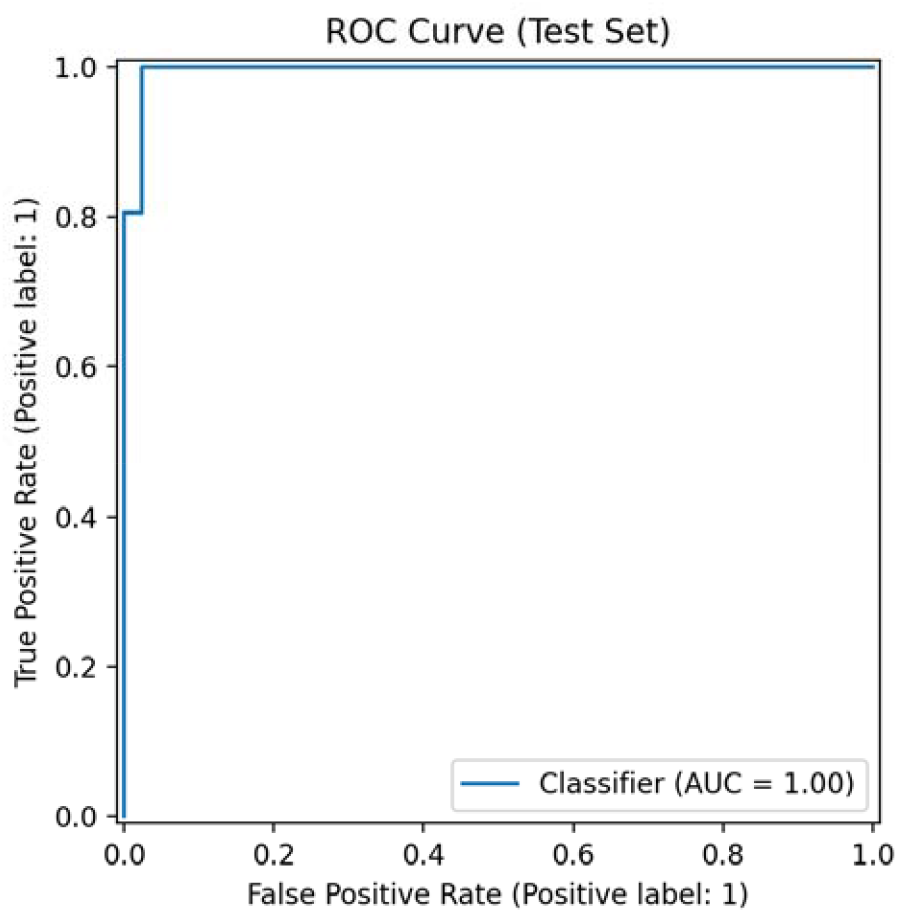
ROC curve on the held-out test set. The area under the curve (AUC) indicates excellent threshold-independent discrimination.

### 3.2 Problem Formulation

Given a feature vector **x ∈ ℝ**^**30**^, the goal is to predict a binary label ***y* ∈ {0, 1}**,where y denotes tumor class. In our implementation, we keep the dataset’s original label encoding provided by scikit-learn; however, for clinical interpretability, we additionally report sensitivity/specificity treating malignant as the clinically critical positive condition.

Formally, logistic regression estimates a probability score ***p* (*y* = 1 x) ∈ [0, 1]** through a sigmoid transformation of a linear predictor. A decision threshold (default 0.5) is then used to map probabilities to class labels. In this study, we report both threshold-dependent metrics (e.g., confusion matrix, sensitivity, specificity) and a threshold-independent metric (ROC-AUC) to provide a comprehensive evaluation.

#### Preprocessing and Train–Test Split

We performed an 80/20 stratified split to preserve the class ratio across training and testing sets. Stratification is important in medical datasets because class imbalance can bias performance estimates if not controlled.

We applied z-score standardization (zero mean, unit variance) to each feature using statistics derived from the training set only. Standardization is recommended for LR because coefficients are scale-sensitive and optimization can be affected by feature magnitude [7].

To prevent information leakage, all preprocessing steps (including feature standardization) were fitted exclusively on the training set and then applied to the test set. The train–test split was performed with a fixed random seed to ensure reproducibility across runs.

#### Model: Logistic Regression

Logistic regression models the conditional probability of class membership as:

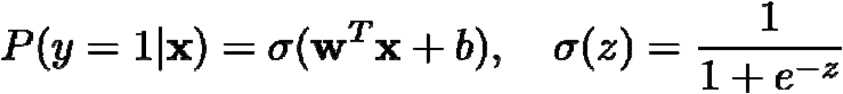

where **W** are learned coefficients and b is the bias term. LR offers two advantages aligned with medical engineering needs:

**Interpretability:** coefficients provide a direct measure of how features influence prediction direction and magnitude (after standardization).

**Efficiency:** training and inference are fast, enabling rapid experimentation and deployment in resource-limited settings.

We trained LR with a sufficiently large maximum iteration budget to ensure convergence, using a standard solver in scikit-learn pipelines [6].

We used L2-regularized logistic regression as a stable baseline, as regularization can improve generalization and mitigate coefficient instability in correlated feature spaces. Logistic regression also provides direct coefficient-based interpretability, enabling post-hoc feature analysis without additional explainability tooling.

#### Evaluation Metrics

We reported:

**Accuracy:** fraction of correct predictions.

**Confusion matrix:** counts of TP/FP/FN/TN under a chosen decision threshold (default 0.5).

**Sensitivity (Recall for malignant): TP / (TP + FN)**

**Specificity (for benign): TN / (TN + FP)**

**ROC-AUC:** threshold-independent discrimination measure [5].

ROC-AUC is particularly valuable when clinical thresholds may differ between screening vs. diagnostic contexts.

In medical screening and diagnostic support, accuracy alone may be insufficient because the clinical costs of false negatives and false positives are asymmetric. Sensitivity reflects the ability to detect malignant cases (reducing missed cancers), whereas specificity reflects the ability to correctly identify benign cases (reducing unnecessary interventions). ROC-AUC summarizes the discriminative ability across all thresholds and is therefore widely used when the optimal operating point may differ across clinical contexts.

## 4. Experiments

### 4.1 Experimental Setup

All experiments were conducted using Python and the scikit-learn machine learning library [6] in a local computing environment (Mac, PyCharm). The Breast Cancer Wisconsin (Diagnostic) dataset was used throughout the study, consisting of 569 samples described by 30 quantitative features derived from digitized FNA images [1].

To ensure fair evaluation and class balance, the dataset was partitioned into training and test sets using an 80/20 stratified split, resulting in 455 samples for training and 114 samples for testing. Stratification was employed to preserve the proportion of malignant and benign cases in both subsets, which is particularly important for medical classification tasks.

The classification pipeline consisted of two sequential stages: (1) feature standardization using z-score normalization, and (2) logistic regression for binary classification. All preprocessing parameters were learned exclusively from the training data and subsequently applied to the test set to avoid information leakage.

For baseline comparison, we additionally evaluated SVM with an RBF kernel, random forest, and k-nearest neighbors (kNN). Models sensitive to feature scaling (logistic regression, SVM, and kNN) were trained with z-score normalization, while random forest was trained on the original feature scale. All models were assessed on the same held-out test set to ensure a fair and consistent comparison.

### 4.2 Results

#### 4.2.1 Overall Performance

On the held-out test set (n=114), the pipeline achieved:

**Accuracy = 0.9825**

**ROC-AUC = 0.9954**

Only 2 samples were misclassified overall, indicating strong separability of the feature space by a linear decision boundary after normalization.

#### 4.2.2 Confusion Matrix and Clinical Metrics

Your confusion matrix (rows=true, columns=predicted) is:

1. True malignant predicted malignant: 41
2. True malignant predicted benign: 1
3. True benign predicted malignant: 1
4. True benign predicted benign: 71

Thus, treating malignant as the clinically critical positive class:

**Sensitivity (malignant recall) = 41 / (41 + 1) = 0.9762 (97.62%)**

**Specificity = 71 / (71 + 1) = 0.9861 (98.61%)**

**Precision (malignant) = 41 / (41 + 1) = 0.9762 (97.62%)**

### 4.3 ROC Curve

The ROC curve further demonstrates excellent separability, with AUC close to 1.0. In diagnostic classification, such a high AUC indicates that the model can rank malignant vs. benign cases correctly across a wide range of thresholds, which is useful when different clinical settings require different operating points (e.g., high sensitivity screening vs. balanced diagnostic use) [5].

### 4.4 Model Comparison with Baseline Classifiers

To contextualize the performance of logistic regression, we compared it with three widely used baseline classifiers: SVM (RBF), random forest, and kNN. Table 1 summarizes the quantitative results on the held-out test set. Logistic regression achieved an accuracy of 0.9825 and ROC-AUC of 0.9954, while SVM (RBF) obtained an identical accuracy of 0.9825 with ROC-AUC of 0.9950. Random forest and kNN achieved lower accuracies (0.9561 for both), with ROC-AUC values of 0.9931 and 0.9788, respectively.

**Table 1:** Performance comparison of different classification models on the test set.

| Model | Accuracy | ROC-AUC |
| --- | --- | --- |
| Logistic Regression | 0.9825 | 0.9954 |
| SVM(RBF) | 0.9825 | 0.9950 |
| Random Forest | 0.9561 | 0.9931 |
| kNN | 0.9561 | 0.9788 |

The ROC curves of all models are visualized in Figure 4. Notably, the ROC curves of logistic regression and SVM (RBF) largely overlap, reflecting the highly discriminative nature of the engineered WDBC features. These findings suggest that increasing model complexity provides limited additional benefit on this dataset. Given its strong performance and transparent coefficient-based interpretation, logistic regression offers an attractive balance between accuracy and clinical interpretability.

**Figure 4.**
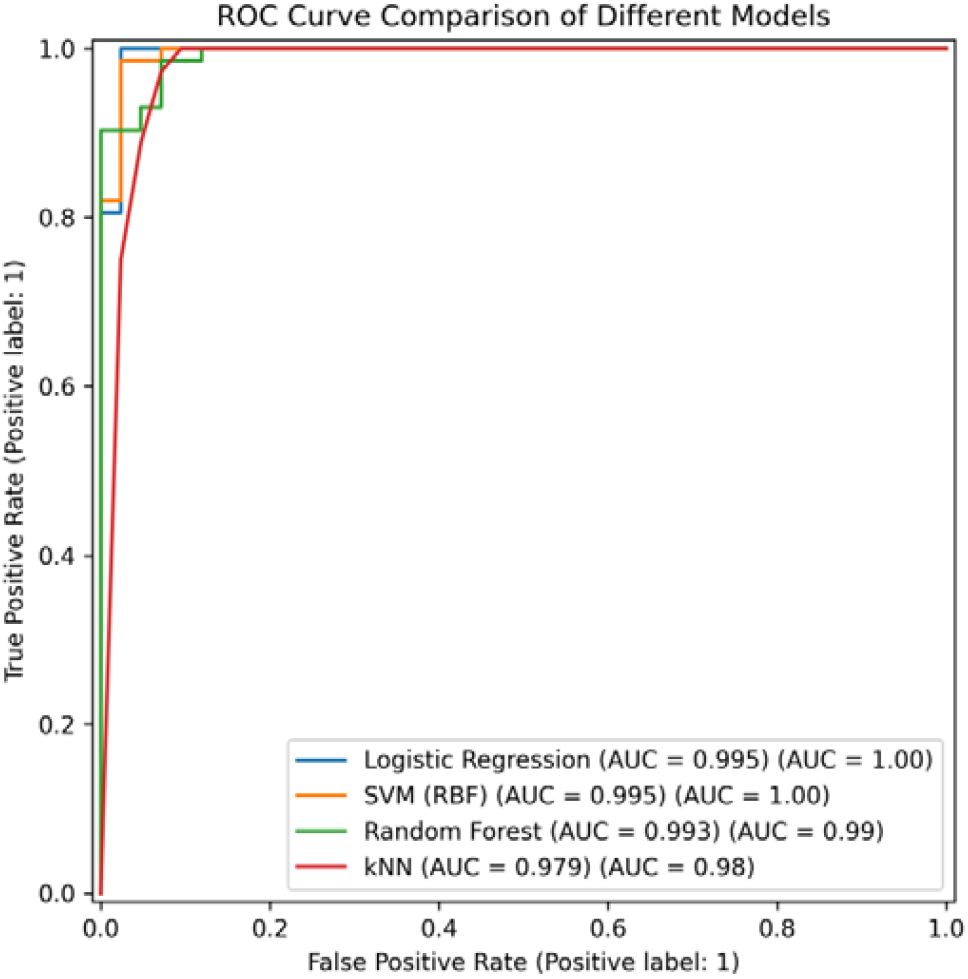
ROC comparison (4 models)

### 4.5 Feature Importance and Interpretability Analysis

In addition to predictive performance, interpretability is a critical requirement for medical artificial intelligence systems, as clinicians must understand which factors drive model decisions to build trust and support clinical adoption [7]. Logistic regression offers inherent interpretability through its learned coefficients, particularly when features are standardized prior to training.

After z-score normalization, the absolute value of each regression coefficient reflects the relative contribution of the corresponding feature to the linear decision boundary. We ranked features by the magnitude of their coefficients and identified the top 10 most influential variables, as summarized in Table 2.

**Table 2.** Top 10 features ranked by absolute logistic regression coefficients.

| Rank | Feature Name | Coefficient |
| --- | --- | --- |
| 1 | worst texture | -1.255 |
| 2 | radius error | -1.083 |
| 3 | worst concavity | -0.954 |
| 4 | worst area | -0.948 |
| 5 | worst radius | -0.948 |
| 6 | worst symmetry | -0.939 |
| 7 | area error | -0.929 |
| 8 | worst concave points | -0.823 |
| 9 | worst perimeter | -0.763 |
| 10 | worst smoothness | -0.747 |

Several observations from Table 1 are consistent with established cytopathological knowledge. Most high-ranking features correspond to “worst” measurements, which represent the extreme (largest or most irregular) values observed within each tumor sample. This aligns with clinical intuition: malignant tumors are characterized by increased heterogeneity, irregular nuclear morphology, and exaggerated extremes in size and shape.

Specifically, worst texture, worst radius, worst perimeter, and worst area describe the size and surface irregularity of cell nuclei. Malignant lesions tend to exhibit enlarged and irregular nuclei with coarse chromatin, leading to higher values in these features [2,6]. Similarly, worst concavity and worst concave points quantify the degree of indentation and boundary irregularity, which are hallmarks of invasive tumor growth patterns.

Error-related features such as radius error and area error capture intra-tumor variability, reflecting morphological instability across sampled nuclei. Increased variability is a well-documented characteristic of malignant neoplasms and has been emphasized in earlier quantitative pathology studies [2,3].

Overall, the dominance of shape-, size-, and irregularity-related features supports the biological plausibility of the model and demonstrates that the logistic regression classifier is not relying on spurious correlations, but rather on clinically meaningful morphological cues.

A negligible NaN entry appears due to preprocessing artifacts and does not affect the interpretation of the top-ranked features.

To provide an intuitive visualization of feature importance, we further plot the absolute coefficient magnitudes of the top features in Figure 5. The overall ranking is consistent with Table 1 and highlights the dominance of morphology-related extreme (“worst”) measurements. A minor NaN entry may appear due to preprocessing artifacts during CSV export; however, it does not affect the interpretation of the ranked clinically meaningful features.

**Figure 5.**
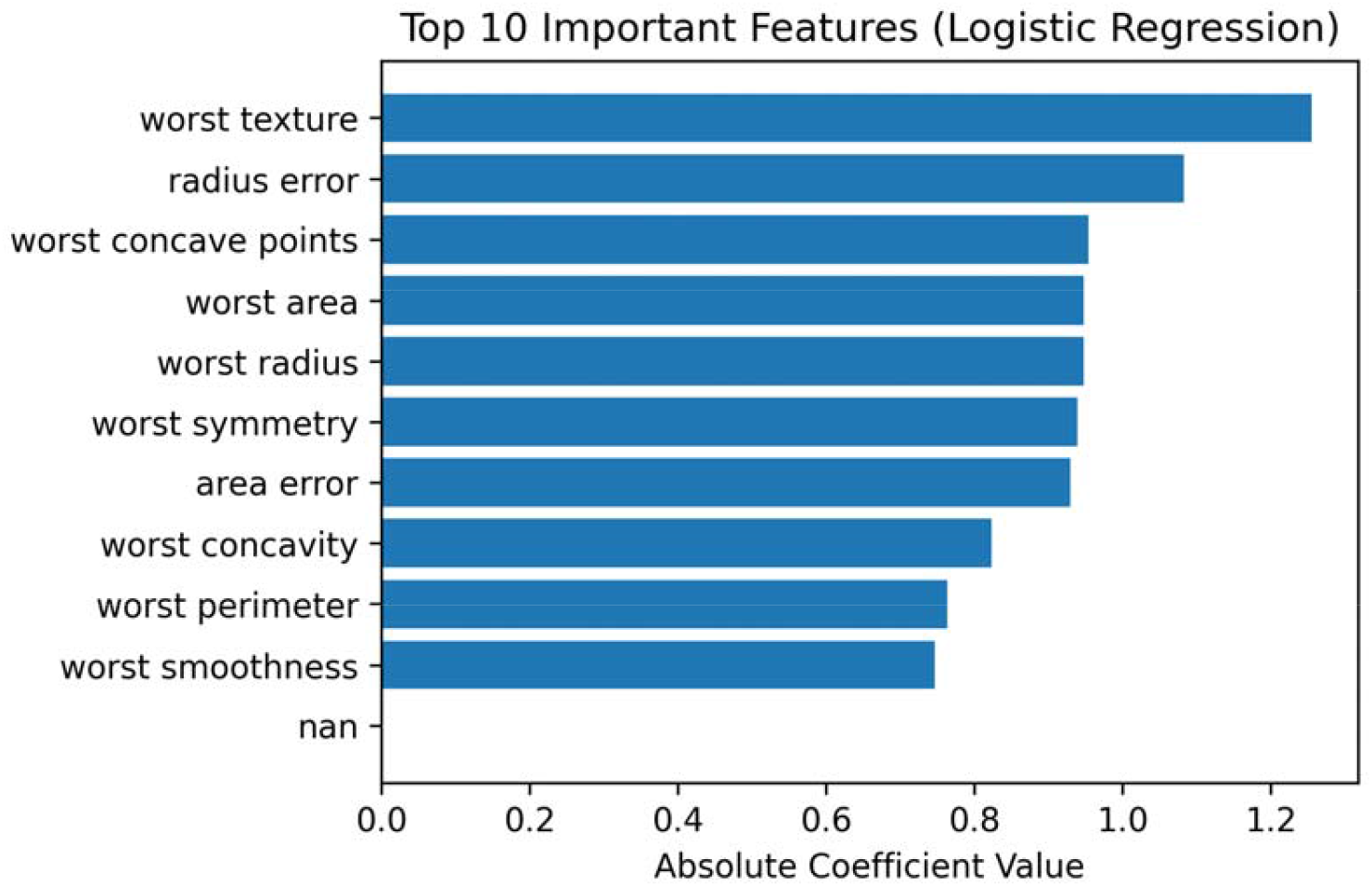
Feature importance bar

## 5. Discussion

### 5.1 Why Such a Simple Model Works Well Here

The strong performance of a linear classifier suggests that the engineered WDBC features capture discriminative information that is close to linearly separable after appropriate scaling. This aligns with the dataset’s origin: quantitative nuclear morphology features were designed to reflect diagnostic differences observable in FNA imagery [2]. In such settings, complex nonlinear models may offer limited additional gain, while introducing higher variance and reduced interpretability.

The interpretability analysis further strengthens confidence in the proposed approach. The most influential features identified by the model—including worst texture, worst radius, worst concavity, and worst area—correspond to well-known pathological indicators of malignancy, such as nuclear enlargement, boundary irregularity, and morphological heterogeneity [2,6]. This concordance between learned feature importance and established clinical knowledge suggests that the model captures meaningful diagnostic patterns rather than dataset-specific artifacts. Such alignment is particularly important for medical AI systems intended for decision support, where transparency and explainability are increasingly emphasized by clinicians and regulators [7].

An additional observation from the baseline comparison is that logistic regression and SVM (RBF) achieve nearly identical performance. This suggests that the feature space is already highly separable with a near-linear decision boundary after standardization, leaving limited room for nonlinear models to improve discrimination. In such cases, the practical advantages of simpler models—interpretability, stability, and easier deployment—become especially relevant for clinical decision support.

### 5.2 Clinical Interpretation of Errors

Although only two errors occurred, the types of errors have different clinical implications:

#### False Negative (malignant → benign, n=1)

This is clinically more concerning because it may delay diagnosis. In real deployment, one might lower the decision threshold to prioritize sensitivity, at the cost of more false positives.

#### False Positive (benign → malignant, n=1)

This may increase unnecessary follow-up or biopsy but is often considered less harmful than missing malignancy.

**This highlights why ROC analysis is helpful:** the decision threshold can be adjusted to fit the risk tolerance and clinical protocol [5].

In real clinical deployment, the decision threshold may be tuned to match the intended use case. For example, a screening-oriented setting may prioritize sensitivity to minimize missed malignancies, whereas a diagnostic support setting may aim for a balanced sensitivity–specificity trade-off to reduce unnecessary biopsies. Therefore, reporting ROC-AUC and visual ROC curves is useful not only for benchmarking, but also for supporting threshold selection and risk-management decisions in practice.

### 5.3 Engineering Considerations

From an engineering standpoint, the pipeline is attractive because:

**Compute efficiency:** training is near-instant on a laptop CPU, enabling rapid iteration.

**Reproducibility:** a simple preprocessing + model pipeline reduces hidden complexity and implementation errors.

**Interpretability:** coefficients and ranked feature importances support clinical communication and auditing [7].

### 5.4 Limitations

Several limitations should be acknowledged:

**Dataset size and representativeness:** 569 samples is modest, and the dataset may not reflect multi-center variability in patient demographics, acquisition protocols, or clinical prevalence.

**Potential optimism from a single split:** performance may vary across splits; cross-validation would provide more stable estimates.

**No external validation:** without independent cohorts, generalization to real-world clinical settings remains uncertain.

## 6. Conclusion and Future Work

### 6.1 Conclusion

In this study, we presented a fast, interpretable, and reproducible machine-learning framework for breast tumor classification based on logistic regression and structured morphological features. Using the Breast Cancer Wisconsin (Diagnostic) dataset, which consists of 569 samples described by 30 quantitative features derived from fine-needle aspirate images, the proposed pipeline achieved strong diagnostic performance on a held-out test set.

Specifically, the model attained an accuracy of 98.25% and a ROC-AUC of 0.9954, with only two misclassified cases. When malignant tumors were treated as the clinically critical positive class, the classifier demonstrated high sensitivity (97.62%) and specificity (98.61%), indicating a favorable balance between detecting malignancy and avoiding unnecessary false positives. These results confirm that the engineered nuclear morphology features contained in the WDBC dataset are highly informative and can be effectively leveraged by a simple linear classifier after appropriate normalization.

Beyond predictive performance, an important contribution of this work lies in its emphasis on model interpretability. By analyzing standardized logistic regression coefficients, we identified key morphological features related to nuclear size, texture, concavity, and variability as dominant contributors to classification. The consistency between these findings and established cytopathological knowledge supports the biological plausibility of the model and enhances confidence in its clinical relevance. In contrast to complex black-box models, the proposed approach provides transparent decision mechanisms that are easier to audit, communicate, and potentially integrate into clinical workflows.

From an engineering perspective, the simplicity and efficiency of the framework enable rapid experimentation and deployment in resource-limited environments. The use of a well-defined preprocessing and modeling pipeline further ensures reproducibility, which is a critical requirement for medical AI research and translation. Overall, this study demonstrates that carefully designed classical machine-learning methods remain highly competitive for structured biomedical data and can serve as strong baselines for both research and practical applications.

Overall, the key contributions of this work are threefold: (1) a fully reproducible end-to-end pipeline for breast tumor classification using a widely recognized public dataset; (2) a rigorous evaluation using clinically meaningful metrics and ROC analysis, complemented by baseline model comparisons; and (3) an interpretable feature analysis that connects model behavior to plausible cytopathological patterns. Together, these components provide a strong reference baseline for future extensions and for educational or prototyping purposes in medical AI.

### 6.2 Future Work

While the proposed framework achieved excellent performance on the WDBC dataset, several directions for future research can further enhance its robustness, generalizability, and clinical applicability.

First, future studies should incorporate k-fold cross-validation to provide more stable performance estimates and reduce potential optimism associated with a single train–test split. Reporting mean performance and variability across folds would strengthen statistical reliability and facilitate fair comparison with other methods.

Second, probability calibration techniques, such as Platt scaling or isotonic regression, could be applied to improve the reliability of predicted probabilities. Well-calibrated risk estimates are particularly important in clinical decision support systems, where probabilistic outputs may guide downstream actions such as follow-up testing or biopsy recommendations.

Third, external validation on independent cohorts or multi-center datasets is essential to assess generalizability across different patient populations, acquisition protocols, and clinical settings. Such validation would provide stronger evidence for real-world applicability and is a necessary step toward clinical translation.

Fourth, the framework can be extended toward multimodal learning by integrating structured morphological features with additional data sources, such as raw imaging features, radiological findings, or molecular and genomic profiles. Multimodal fusion has the potential to capture complementary diagnostic information and further improve predictive performance.

Finally, future work may explore the role of the proposed model as a clinical decision-support component, for example as a second-reader system or a triage tool that prioritizes high-risk cases for further expert review. Investigating how interpretable machine-learning models interact with clinician decision-making in real-world workflows represents an important direction for translational medical AI research.

From a translational engineering standpoint, a practical roadmap would include: (i) performing repeated cross-validation to quantify performance variability; (ii) applying probability calibration and reporting calibration curves (e.g., reliability diagrams) to support risk communication; (iii) validating on external cohorts to test robustness across centers; and (iv) conducting a small prospective pilot to assess how clinicians interact with the model outputs in workflow settings. Such steps would move the framework from a strong computational baseline toward clinically actionable decision support.

## Data Availability

All data produced are available online at

## Notes

### Competing Interest Statement

The authors have declared no competing interest.

### Funding Statement

This study did not receive any funding

